# Clinically relevant shifts in endogenous and exogenous mutational processes proximate to metastasis support local consolidative treatment in EGFR-driven non-small cell lung cancer

**DOI:** 10.1101/2021.05.04.21256425

**Authors:** J. Nicholas Fisk, Amandeep R. Mahal, Alex Dornburg, Stephen G. Gaffney, Sanjay Aneja, Joseph N. Contessa, David Rimm, James B. Yu, Jeffrey P. Townsend

## Abstract

The progression of cancer—including the acquisition of therapeutic resistance and the fatal metastatic spread of therapy-resistant cell populations—is an evolutionary process that is challenging to monitor between sampling timepoints. Here we apply mutational signature analysis to clinically correlated cancer chronograms to detect and describe the shifting mutational processes caused by both endogenous (e.g. mutator mutation) and exogenous (e.g. therapeutic) factors between tumor sampling timepoints. In one patient, we find that cisplatin therapy can introduce mutations that increase the likelihood of genetic adaptation to subsequent targeted therapeutics. In another patient, we trace the emergence of known driver mutation CTNNB1 S37C to specific detection of defective mismatch repair associated mutational signature SBS3. Metastatic lineages were found to emerge from a single ancestral lineage arising during therapy—a finding that argues for the consideration of local consolidative therapy over other therapeutic approaches in EGFR-positive non-small cell lung cancer. Broadly, these results demonstrate the utility of phylogenetic analysis that incorporates clinical time course and mutational signature detection to inform clinical decision making and retrospective assessment of disease etiology.

## Introduction

Cancer progression is an evolutionary process, enabling tumors to evade our best therapeutics and, ultimately, to recur and metastasize. The majority of cancer mortality occurs through the metastatic spread of therapy-resistant cell populations^1^. The evolution of therapeutic resistance *in vivo* remains enigmatic in part because it must be reconstructed from patient biopsies whose timing and location are dictated by patient care, rather than from design to empower discovery. This limitation prevents direct observation of cancer’s evolution between clinically-appropriate sampling timepoints and limits investigation of tumor evolution in response to medical intervention. By understanding the genetic evolution of clinically-important characteristics within these incidental sampling intervals, we can potentially gain insight into both the personal and the general etiology of disease progression.

First-line targeted therapies for epidermal growth factor receptor (EGFR)-positive Non-Small Cell Lung Cancer (NSCLC) offer excellent, initially sustained clinical response. Following initial treatment, patients are at risk of developing delayed metastatic disease^2^, which can be precipitated by the common EGFR T790M mutation. EGFR T790M arises within a tumor due to two distinct evolutionary processes: mutation, which leads to its appearance in single cells of the tumor, and selection, which increases its frequency in the tumor in response to treatment. Several *in vitro* experiments have suggested EGFR-inhibitor resistance arises due to strong selection during treatment^3–5^. However, determining the cause of the mutation and whether it arises in a single ancestral cell lineage before a metastatic cascade or separately among multiple metastatic lineages remains uncertain. Patient care seldom takes the form of a single, monolithic intervention. A better understanding could circumvent clinical trajectories that increase the frequency of resistance, or support pursuit of local consolidative treatment (LCT) over alternative approaches such as maintenance therapy^6^.

Here, we use techniques from evolutionary biology that reconstruct ancestral states of genetic mutations to infer relationships among clonal lineages and reveal temporal patterns of divergent events in metastatic progression^7^. We combine Bayesian inference of cancer chronograms with mutational signature analysis to deeply probe tumor evolution in two patients with metastatic lung adenocarcinoma, providing the first-in-human, clinically correlated tumor phylogenetic analysis of the metastatic cascade in EGFR-mutated NSCLC. Through examination of the phylogenetic topology and branchwise mutational signature analysis, we contextualize the effect of known endogenous and exogenous mutational processes within the evolution of individual patient metastatic cascades. Specifically, we investigate whether exogenous exposure to platinum therapy preceding erlotinib could precipitate T790M mutation. We then examine whether endogenous mutational processes could play a similar role introducing new mutations into a dynamically evolving tumor. Lastly, we test whether such mutations arising in therapy-associated tumor lineage bottlenecks lead to single lineages that form the basis of multiple therapy-resistant metastatic events. We use these results to evaluate the clinical relevance of shifts in endogenous and exogenous mutational processes and to motivate clinical consideration of potential opportunities for local consolidative treatment.

## Materials and Methods

### Tumor Sampling and Sequencing

The tissues assessed in this study were obtained from the Yale Pathology Archives based on Yale Human Investigation Committee at Yale University, Protocol no. 0304025173 to Dr. David L. Rimm, which enabled retrieval of tissue from archives that was consented or had been approved for use with waiver of consent. Sample collection and tumor sequencing were performed as described in Zhao et al^8^. Clinical information was retrospectively obtained for this investigation from electronic medical records after approval from the Yale University Human Investigation Committee (HIC 1508016314).

### Variant Calling

Variant calling was performed using the protocol described in Zhao et al.^8^ using the same, but updated, SNP reference databases (accessed July 2019). Briefly this approach involves alignment of whole-exome capture Illumina HiSeq reads to the hg19 human reference genome using Eland. Somatic single-nucleotide variants are then provisionally called using a combination of GATK and freebayes, filtering out polymorphisms within the National Heart, Lung, and Blood Institute Exome Sequencing Project, 1000 Genomes, or Yale Human Exome Database. These somatic variants are then analyzed for false negatives using the multinomial approach also outlined in Zhao et al.^8^

### Phylogenetic Analysis

Evolutionary relationships and divergence times were simultaneously estimated in BEAST v.2.5.2 ^9^ assuming an uncorrelated model of molecular rates with a lognormal distribution (UCLN) and a coalescent exponential population growth branching process prior. Divergence times were calibrated using 1) a lower bound for the most recent common ancestor of the germline lineages at the patients’ year of birth, and 2) a non-contemporaneous tip age for primary tumor sequences at the time of biopsy and contemporaneous tip ages for all metastatic samples taken during autopsy. Each cancer chronogram was inferred by three independent runs lasting 5.0 × 10^8^ iterations, sampling every 1,000 generations. Sufficient sampling of the posterior distribution for each parameter was evaluated via computation of effective sample size (ESS) values, with ESS values greater than 200 indicating adequate sampling of a target parameter. Independent runs were then assessed for convergence and appropriate levels of burn-in through visual inspection of the marginal posterior probabilities versus the generation state using Tracer v.1.6. Sampled posteriors from these three consistent executions were combined with TreeAnnotator v.2.4 to generate a maximum clade-credibility tree that summarized the posterior distribution of estimated evolutionary relationships and branch lengths.

To ensure the robustness of the tumor tree topology to the choice of alternate phylogenetic inference approaches, we validated the tree topology using maximum-likelihood in IQ-TREE^10^. We conducted 1000 bootstrap iterations of a GTR model on the alignment and found that the bootstrap support corresponded to the strength of topology indicated by the Bayesian distributions. Ancestral states of variant nucleotides were then reconstructed via maximum likelihood in FastML^11^ using the maximum clade-credibility tree under a GTR substitution model specifying a gamma distribution of rates discretized into eight categories. These ancestral states were found to be entirely consistent with those obtained by computing the ancestral states using the empirical-Bayes method in IQ-TREE and when taking the average probability determined by FastML across sampled posterior tree states.

### Tracing of Mutational Processes Associated with COSMIC Mutational Signatures

Mutational signatures were assessed using deconstructSigs (v.1.9.0) ^12^, referencing only lung-associated endogenous and any known exogenous signatures (COSMIC Exome reference signatures, version 3, May 2019) and enforcing a standard minimum signature contribution threshold of 0.05 and ensuring cosine-similarity between called signatures does not exceed COSMIC’s own. The proportions of mutations attributable to these mutational signatures were traced along patient chronograms for each set of inferred ancestral-state variants along the branches of the phylogeny.

### Code and Data Availability

Input files for phylogenetic analyses, as well as other code, intermediate files, and diagnostic images are available at https://github.com/Townsend-Lab-Yale/LUAD-PhyLCT. The sequencing reads have been made publicly available through the NCBI Sequence Read Archive, published under the BioProject accession PRJNA674368 on November 3rd, 2020.

## Results

### Platinum Therapy Preceding Erlotinib Therapy Precipitates T790M Mutation

We applied our Bayesian phylogenetic inference and mutational signature analysis to two patients. Patient 435 received cisplatin, a platinum-based chemotherapeutic, coupled with pemetrexed (Pe), shortly after primary tumor biopsy, then over a year later received the EGFR tyrosine kinase inhibitor erlotinib. Our phylogenetically informed mutational signature inference (Figure 1) illustrates that the known platinum-associated mutational signature SBS35 (light grey) is coincident with cisplatin therapy: there is no SBS35 signature attributed to the ancestor of the primary tumor and metastatic lineages, and the highest proportion of mutations attributable to this signature (19.1%) is observed in the inferred ancestor immediately subsequent to cisplatin exposure. The EGFR T790M resistance mutation is coincident with this peak proportion of cisplatin mutations, occurring along the same branch in the phylogeny via the nucleotide substitution c.2369C→T. Signature SBS35 is especially associated with C→T mutations, which constitute 20.3% of mutations associated with the signature. Preceding erlotinib therapy with cisplatin therapy presumably reduces the cancer cell population size, but simultaneously induces specific genetic heterogeneity that potentially includes initially undetectable low-frequency cancer cells with the T790M mutation. While signature SBS35 persisted until end-of-life, the cessation of cisplatin therapy led to serially smaller average proportions (≤12.4%) of mutations attributable to SBS35 across the entire metastatic clade, as mutations arising from other processes increased while mutations from SBS35 had already plateaued—as early as three years prior to sampling of the metastases at autopsy.

**Figure 1:**
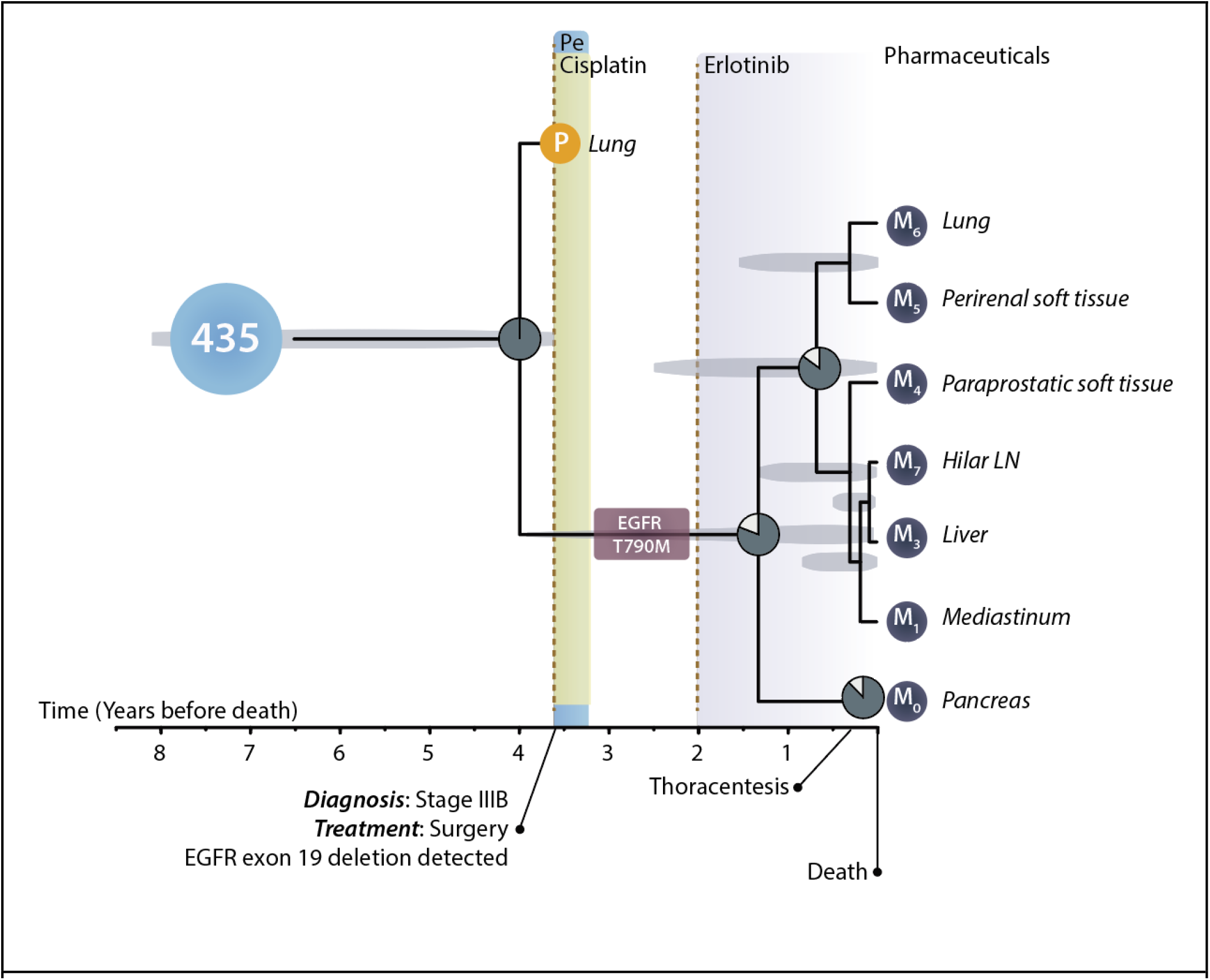
Tumor chronogram reflecting the temporal evolution of primary and metastatic tissues in relation to disease progression within patient 435 (normal tissue, light blue circle), a man incidentally diagnosed with stage-IIIB lung cancer on routine chest x-ray at 70-80 years of age, just over 3.5 years before death (BD). The most recent common ancestor of the primary tumor (yellow circle) and all metastases was reconstructed at 4 years before death (BD). Resection of the primary tumor of the lung (yellow circle) was attempted upon diagnosis just over 3.5 years BD, and initial pathology revealed an EGFR Exon-19 deletion. The patient underwent postoperative chemotherapy with pemetrexed (blue shading) and cisplatin (green shading). EGFR-targeted therapy had yet to be FDA approved. Proportions of mutations attributable to cisplatin-associated SBS35 (white slice) and to other signatures (dark grey pie) are indicated at ancestral nodes in the tumor tree. Fourteen months (1.2 years) after his last cycle of pemetrexed and cisplatin, his disease progressed, and he then received the EGFR-targeted therapy erlotinib (purple shading). EGFR T790M mutation arose to detectable frequency some time between 1.5–4 years BD (maroon box), presumably originating at low frequency during cisplatin therapy between 3–3.5 years and rising in frequency in response to treatment with erlotinib 1.5–2 years BD. Twenty months (1.7 years) after initiation of erlotinib, his disease continued to progress, and he developed a malignant pleural effusion. He died of progressive metastatic disease. Metastatic tissue (dark blue circles) was sampled at autopsy from his hilar lymph nodes, liver, lungs, mediastinum, pancreas, as well as perirenal and para-prostatic soft tissue. The length of the truncal branch is truncated so as to provide sufficient space to display treatment, progression, and tumor tree. Light-grey ovals underlying each ancestral node indicate the 95% highest posterior density interval for each node age estimate.

### CTNNB1 S37C Mutation Follows Treatment with Bevacizumab and Precipitates Defective Mismatch-Repair

We conducted tumor-tree inference and mutational signature analysis on a second patient. This patient received first-line erlotinib therapy before disease progression at 1 year, prompting a switch to carboplatin, pemetrexed, and bevacizumab. Following the onset of bevacizumab treatment, a lineage diverged and gave rise to a highly vascular splenic metastatic mass and a distinct metastatic clade with an epithelial-mesenchymal transition- and angiogenesis-associated CTNNB1 S37C mutation. Less than a year before the time of death, lineages of this CTNNB1-mutant metastatic clade diverged genetically and colonized adrenal, lung, kidney, paratracheal lymph node, and liver tissues (**Fig. 2**). The known defective mismatch repair mutational signature SBS3 (light grey) followed acquisition of CTNNB1 S37C: there is no detectable SBS3 signature on any lineage lacking the mutation (**Fig. 2**), including the contemporaneous lesion in the highly vascularized spleen. Unlike the exogenously generated SBS35 signature in patient 435— which attenuated after discontinuation of cisplatin therapy (**Fig. 1**)—the endogenously generated SBS3 signature only becomes more prominent over time subsequent to the clonal growth of the CTNNB1 S37C lineage and its metastatic lineage diversification. Only 7.8% of mutations in the common ancestor of the non-splenic metastases were attributable to signature SBS3. In contrast, 17.8% and 16.9% of the attributable mutations in the liver lesion and the lineage leading to the other CTNNB1 S37C metastases were attributable to signature SBS3. This signature of DNA mismatch repair deficiency is specific to the metastatic lineages that arose subsequent to the appearance of the CTNNB1 S37C mutation.

**Figure 2:**
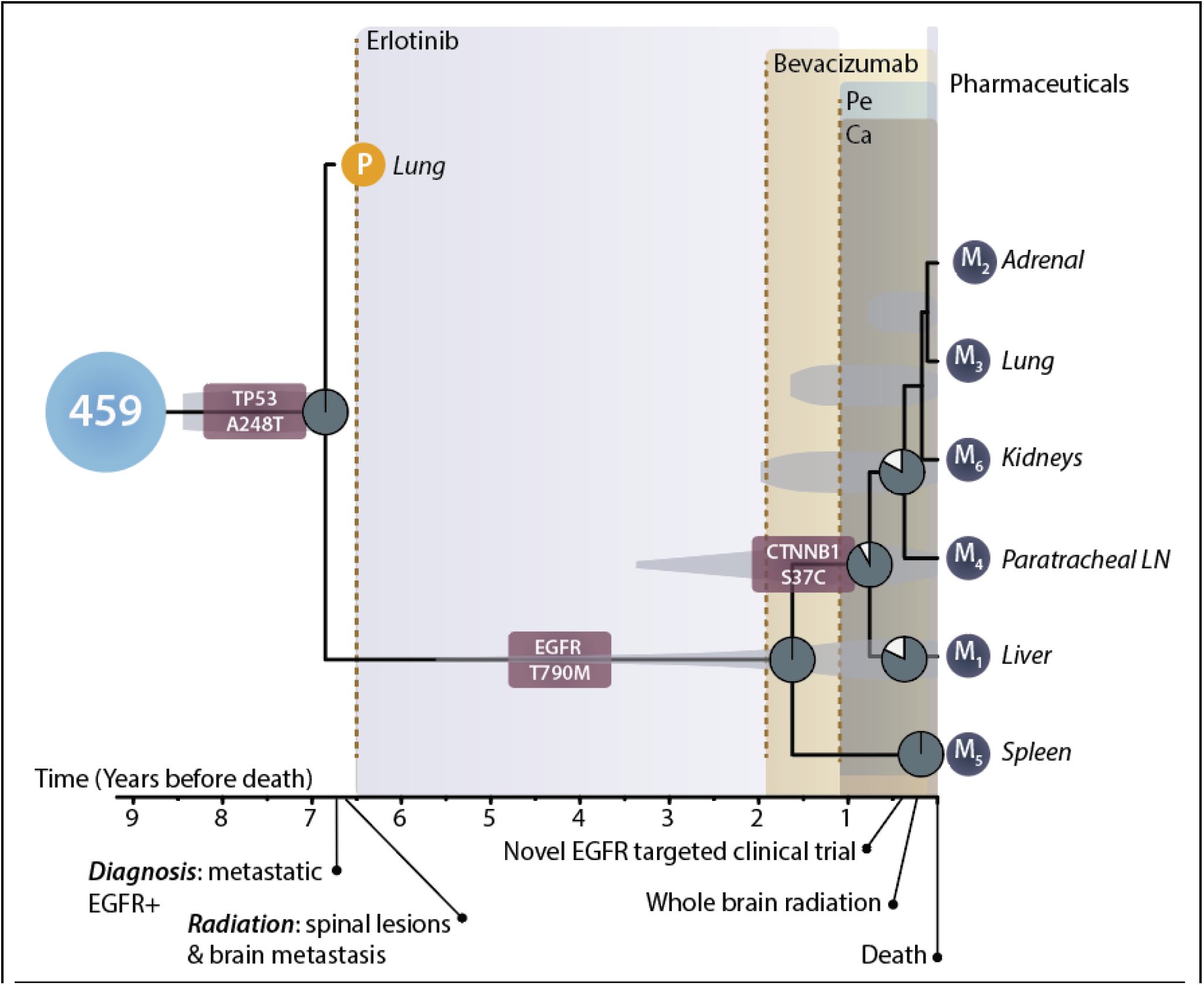
Tumor chronogram reflecting the temporal evolution of primary and metastatic tissues in relation to disease progression within patient 459 (normal tissue, light blue circle), a woman diagnosed at 45-50 years of age with metastatic lung cancer at initial presentation. A most recent common ancestor of the primary tumor and all metastases was reconstructed at 6.85 years before death (BD) and featured a TP53 mutation (maroon rounded rectangle). She subsequently underwent palliative radiotherapy to the spine and brain before then receiving EGFR-targeted therapy (erlotinib, purple shading). Fifty-four months (4.5 years) after initiation of erlotinib, her disease progressed, and her therapeutic regimen was expanded to include bevacizumab (beige shading) in addition to continued erlotinib.One year after beginning bevacizumab, the disease progressed further and therapy was switched to carboplatin (brown shading), pemetrexed (green shading), and bevacizumab (yellow shading), with discontinuation of erlotinib treatment. EGFR T790M mutation (maroon rounded rectangle) arose to detectable frequency some time between 1.6–6.5 years BD. A CTNNB1 mutation (marron rounded rectangle) similarly reached detectable frequency in the non-splenic lesions during bevacizumab therapy. Proportions of mutations attributable to defective homologous recombination-based DNA damage repair signature SBS3 (white slice) and attributable to other signatures (dark blue pie) are indicated at ancestral nodes in the tumor tree. Four months BD, the patient underwent therapy with a novel EGFR-targeted compound (AP26113) in a clinical trial due to sustained disease progression. Two months and one month prior to her death, additional treatment with whole-brain radiation and re-initiation of erlotinib were elected, respectively. She died of progressive metastatic disease. Metastatic tissue (dark blue circles) was sampled at autopsy from her adrenal gland, kidney, liver, lungs, paratracheal lymph nodes, and spleen. The length of the truncal branch is truncated so as to provide sufficient space to display treatment, progression, and tumor tree. Light-grey ovals underlying each ancestral node indicate the 95% highest posterior density interval for each node age estimate.

## Discussion

Here we have demonstrated that placing patient clinical treatment data and disease progression into the context of the molecular evolutionary history of tumor lineages provides clinically relevant insights. In one case, we showed how cisplatin-therapy induced a temporally localized burst of mutations, precipitating the rapid evolution of EGFR T790M resistance to erlotinib. In a second case, a CTNNB1 S37C mutation arose during the administration of Bevacizumab, preceding the rapid diversification of tumor lineages with a signature of DNA mismatch repair deficiency and leading to a clade of metastases. In both cases, metastatic lineages were found to emerge from a single ancestral lineage arising during therapy. Collectively, these results demonstrate the clinical relevance of shifts in mutational signatures that result from shifting endogenous and exogenous mutational processes, and support pursuit of local consolidative treatments in EGFR-driven non-small cell lung cancer. Where mutagenic chemotherapy and targeted systemic agents are initial first-line options, traditional chemotherapeutics in the first-line setting may decrease the time to progression associated with targeted agents due to the higher likelihood of extant chemotherapy-induced low-frequency escape mutations.

These two cases illustrate that endogenous and exogenous mutational processes provide the raw material for selective evolution enabling the resistance to therapy. The EGFR T790M mutation is known to convey resistance to erlotinib^13,14^, and our results suggest that CTNNB1 S37C is rescuing a tumor lineage from the anti-angiogenic selection pressure applied by therapy with bevacizumab ^15–17^. However, confirmation of this role necessitates estimation of the strength of selection for resistance or rescue mutations which requires application of cancer effect size estimation^18^ to distinct branches of tumor trees that are contemporaneous with clinical treatment across a larger cohort with samples of normal, primary, and recurrent or metastatic tumor tissue. Future clinical trials or other studies incorporating metachronous sampling that apply such approaches have great potential to illuminate the clinical implications of serial therapeutic treatments on progression.

It might be counterintuitive that a CTNNB1 mutation could be considered a rescue mutation from treatment with bevacizumab when treatment with bevacizumab is clinically indicated by the presence of CTNNB1. However, both bevacizumab and CTNNB1 operate—at least in part—by shifting the tumor along a phenotypic axis of angiogenesis via modulation of the VEGF pathway. CTNNB1 S37C is a putative gain-of-function mutation associated activation of the Wnt pathway, which is associated with loss of mismatch repair function^19–21^. Through its role in Wnt signaling, CTNNB1 also increases VEGF expression by binding its gene promoter^22^—which features seven confirmed consensus binding sites for the beta-catenin/TCF complex—stimulating angiogenesis^23,24^. Treatment with bevacizumab inhibits VEGF function. In the context of a tumor with CTNNB1 mutation, treatment with bevacizumab abrogates the increase in angiogenesis and consequent benefits to tumor growth and proliferation of CTNNB1 mutation. In the context of a CTNNB1 wildtype tumor, bevacizumab decreases angiogenesis, throttling the tumor, and CTNNB1 mutation rescues angiogenesis, benefitting tumor growth and proliferation. Our results lend support to the hypothesis that CTNNB1 S37C can act as a rescue mutation from VEGF-inhibitors.

Patient 459 exhibited no detectable SBS31 or SBS35 signature despite receiving a short end-of-life treatment with carboplatin. It is possible that the signatures of low-frequency mutations may not have been detected by constantly improving machine-learning oriented approaches to deconvolve mutational signatures within tumors^25–31^. However, the presence of SBS35 in all extant and inferred tumor samples subsequent to cisplatin therapy in patient 435 supports the consistency and specificity of signature extraction from tree branches, as does the absence of SBS3 on the early diverging spleen lineage in patient 459, which lacks mutations associated with defective homologous recombination repair. Instead, this lack of immediate detection following treatment is more likely attributable to the lag time before mutations that arise in single cells drift or are selected to clonal fixation^32^. Mutations require time to reach detectable prominence in the tumor via sustained replication and clonal growth.

The observation of a clade of lineages giving rise to metastases with a single common ancestor with the primary tumor implies a strong selective bottleneck associated with erlotinib therapy. Extant low-frequency mutations can form the basis of evolving resistance to targeted therapies, suggesting there is opportunity for local consolidative treatment (LCT) to prevent later metastatic disease. Compared to utilizing first-line treatment in isolation, multi-modality management (such as high-precision ablative radiotherapy or surgery) could alter the trajectory of disease and provide a survival benefit. Notably, recent randomized controlled phase-II trials have confirmed that patients with oligometastatic EGFR-positive NSCLC experience prolonged overall survival and progression-free survival with LCT ^6,33,34^. This prolonged survival suggests that early intervention with LCT to extinguish the reservoir of drug-tolerant cells, and prevent the emergence of resistant subclones, prior to divergence of a metastatic lineage within the primary tumor could result in significant therapeutic benefit. Anticipatory treatment of evolving cancers, such as is provided by early LCT, may improve clinical outcomes. Given the potential for radiation to induce heterogeneous mutations, this work also suggests that ablative radiation doses should be used as less-than-ablative doses may also have the unintended potential to accelerate potential tumor escape from targeted therapy.

The impact of therapeutic intervention and shifts in mutational processes driven by genic aberration can be deconvolved using mutational signature analyses that provide deeper insights into the tumor evolution than the same analyses devoid of phylogenetic information. These insights include several important clinical implications regarding the timing of DNA-damaging therapy and targeted cancer treatment for which acceleration of mutational heterogeneity may be detrimental. The presence of SBS35 subsequent to cisplatin therapy in patient 435 in particular has clinical implications regarding the sequencing of chemotherapeutics that increase mutational loads and targeted treatment. The same can be said for pre-systemic therapy or concurrent radiation treatment, if radiation doses are used that are unlikely to be ablative. Our study serves as a template for future evolutionary studies that seek to uncover the etiology of metastases that are essential steps to understanding general rules that shape the evolutionary trajectory of cancer. Developing an understanding of how dynamic mutational processes evolve and result in the diversification of metastatic lineages can inform clinical decision making, guide the design of treatment paradigms, and improve patient outcomes.

## Data Availability

https://www.ncbi.nlm.nih.gov/bioproject/PRJNA674368

## Author contributions

JNF and ARM contributed equally to this work. JPT and JY conceived of the study. DRM and JNC provided materials and helped with clinical data collection. ARM, SA, and JY reviewed, gathered, and organized patient clinical data. SGG performed initial bioinformatic analysis of sequence data. JNF and AD performed phylogenetic and cancer chronogram inference. JNF performed phylogenetic analyses of mutational signatures. All authors contributed to interpretation of results. JNF, ARM, JPT, and AD drafted the manuscript, with contributions from SGG and JY. All authors approved the final version of the manuscript.

## Data and code availability

All data and code used in the analysis is referenced within the Methods and Supplementary Tables.

## Acknowledgements

JPT and DR acknowledges Gilead Sciences, Inc, for funding supporting this research, as well as the Elihu endowment and the Notsew Orm Sands Foundation. JNF acknowledges NLM Grant 5T15LM007056-32 and NCI Grant 1F31CA257288-01. We also thank the Yale Center for Research Computing for their continued support.

## References

1. Dillekås, H., Rogers, M. S. & Straume, O. Are 90% of deaths from cancer caused by metastases? Cancer Med. 8, 5574–5576 (2019).

2. Stewart, E. L., Tan, S. Z., Liu, G. & Tsao, M.-S. Known and putative mechanisms of resistance to EGFR targeted therapies in NSCLC patients with EGFR mutations-a review. Transl Lung Cancer Res 4, 67–81 (2015).

3. Ortiz-Cuaran, S. et al. Heterogeneous Mechanisms of Primary and Acquired Resistance to Third-Generation EGFR Inhibitors. Clin. Cancer Res. 22, 4837–4847 (2016).

4. Bolan, P. O. et al. Genotype-Fitness Maps of EGFR-Mutant Lung Adenocarcinoma Chart the Evolutionary Landscape of Resistance for Combination Therapy Optimization. Cell Syst 10, 52–65.e7 (2020).

5. Foggetti, G. et al. Genetic determinants of EGFR-Driven Lung Cancer Growth and Therapeutic Response In Vivo. Cancer Discov. (2021) doi:10.1158/2159-8290.CD-20-1385.

6. Gomez, D. R. et al. Local Consolidative Therapy Vs. Maintenance Therapy or Observation for Patients With Oligometastatic Non-Small-Cell Lung Cancer: Long-Term Results of a Multi-Institutional, Phase II, Randomized Study. J. Clin. Oncol. 37, 1558–1565 (2019).

7. Somarelli, J. A. et al. Molecular Biology and Evolution of Cancer: From Discovery to Action. Mol. Biol. Evol. 37, 320–326 (2020).

8. Zhao, Z.-M. et al. Early and multiple origins of metastatic lineages within primary tumors. Proc. Natl. Acad. Sci. U. S. A. 113, 2140–2145 (2016).

9. Bouckaert, R. et al. BEAST 2.5: An advanced software platform for Bayesian evolutionary analysis. PLoS Comput. Biol. 15, e1006650 (2019).

10. Minh, B. Q. et al. IQ-TREE 2: New Models and Efficient Methods for Phylogenetic Inference in the Genomic Era. Mol. Biol. Evol. 37, 1530–1534 (2020).

11. Ashkenazy, H. et al. FastML: a web server for probabilistic reconstruction of ancestral sequences. Nucleic Acids Res. 40, W580–4 (2012).

12. Rosenthal, R., McGranahan, N., Herrero, J., Taylor, B. S. & Swanton, C. DeconstructSigs: delineating mutational processes in single tumors distinguishes DNA repair deficiencies and patterns of carcinoma evolution. Genome Biol. 17, 31 (2016).

13. Pao, W. et al. Acquired Resistance of Lung Adenocarcinomas to Gefitinib or Erlotinib Is Associated with a Second Mutation in the EGFR Kinase Domain. PLoS Med. 2, e73 (2005).

14. Demuth, C. et al. The T790M resistance mutation in EGFR is only found in cfDNA from erlotinib-treated NSCLC patients that harbored an activating EGFR mutation before treatment. BMC Cancer 18, 1–5 (2018).

15. Culy, C. Bevacizumab: antiangiogenic cancer therapy. Drugs Today 41, 23–36 (2005).

16. Haibe, Y. et al. Resistance Mechanisms to Anti-angiogenic Therapies in Cancer. Front. Oncol. 10, (2020).

17. von Felden, J. et al. Mutations in circulating tumor DNA predict primary resistance to systemic therapies in advanced hepatocellular carcinoma. Oncogene 40, 140–151 (2021).

18. Cannataro, V. L., Gaffney, S. G. & Townsend, J. P. Effect Sizes of Somatic Mutations in Cancer. J. Natl. Cancer Inst. 110, 1171–1177 (2018).

19. Chmara, M. et al. Multiple pilomatricomas with somatic CTNNB1 mutations in children with constitutive mismatch repair deficiency. Genes Chromosomes Cancer 52, 656–664 (2013).

20. Serebryannyy, L. A., Yemelyanov, A., Gottardi, C. J. & de Lanerolle, P. Nuclear α- catenin mediates the DNA damage response via β-catenin and nuclear actin. J. Cell Sci. 130, 1717–1729 (2017).

21. Castiglia, D. et al. Concomitant activation of Wnt pathway and loss of mismatch repair function in human melanoma. Genes Chromosomes Cancer 47, 614–624 (2008).

22. Easwaran, V. et al. beta-Catenin regulates vascular endothelial growth factor expression in colon cancer. Cancer Res. 63, 3145–3153 (2003).

23. Pate, K. T. et al. Wnt signaling directs a metabolic program of glycolysis and angiogenesis in colon cancer. EMBO J. 33, 1454–1473 (2014).

24. Olsen, J. J. et al. The Role of Wnt Signalling in Angiogenesis. Clin. Biochem. Rev. 38, 131–142 (2017).

25. Bayati, M. et al. CANCERSIGN: a user-friendly and robust tool for identification and classification of mutational signatures and patterns in cancer genomes. Sci. Rep. 10, 1286 (2020).

26. Afsari, B. et al. Supervised mutational signatures for obesity and other tissue-specific etiological factors in cancer. Elife 10, (2021).

27. Lyu, X., Garret, J., Rätsch, G. & Lehmann, K.-V. Mutational signature learning with supervised negative binomial non-negative matrix factorization. Bioinformatics 36, i154–i160 (2020).

28. Thutkawkorapin, J., Eisfeldt, J., Tham, E. & Nilsson, D. pyCancerSig: subclassifying human cancer with comprehensive single nucleotide, structural and microsatellite mutational signature deconstruction from whole genome sequencing. BMC Bioinformatics 21, 128 (2020).

29. Gulhan, D. C., Lee, J. J.-K., Melloni, G. E. M., Cortés-Ciriano, I. & Park, P. J. Detecting the mutational signature of homologous recombination deficiency in clinical samples. Nat. Genet. 51, 912–919 (2019).

30. Huang, P.-J. et al. mSignatureDB: a database for deciphering mutational signatures in human cancers. Nucleic Acids Res. 46, D964–D970 (2018).

31. Baez-Ortega, A. & Gori, K. Computational approaches for discovery of mutational signatures in cancer. Brief. Bioinform. 20, 77–88 (2019).

32. Foo, J., Leder, K. & Michor, F. Stochastic dynamics of cancer initiation. Phys. Biol. 8, 015002 (2011).

33. Iyengar, P. et al. Consolidative Radiotherapy for Limited Metastatic Non-Small-Cell Lung Cancer: A Phase 2 Randomized Clinical Trial. JAMA Oncol 4, e173501 (2018).

34. Palma, D. A. et al. Stereotactic ablative radiotherapy versus standard of care palliative treatment in patients with oligometastatic cancers (SABR-COMET): a randomised, phase 2, open-label trial. Lancet 393, 2051–2058 (2019).

